# Leadless Pacemaker with Transcatheter Aortic Valve Implantation: A Single Center Experience

**DOI:** 10.1101/2022.06.04.22275979

**Authors:** Feng Gao, Riyad Kherallah, Mackenzie Koetting, Leo Simpson, John Seger, Srikanth Koneru, Joseph Coselli, Ourania Preventza, Vicente Orozco-Sevilla, Nastasya Manon, Guilherme V Silva

**Author notes:** **Corresponding Author** Feng Gao M.D., Internal Medicine, Department of Medicine, Building Tower West McNair Campus (MCHA) A10.193 MS, Houston, TX 77030. **Data availability statement:** The data generated or analyzed in the current study are available from the corresponding author upon request. **Funding:** none. **Ethics approval statement:** This study was approved by the local institutional review board. **Patient consent statement:** Given retrospective nature of study, patient consent was waived with approval from local institutional review board. **Permission to reproduce material from other sources:** All tables and figures were original from study authors. **Clinical trial registration:** not applicable. **Author financial disclosures and emails:** Feng Gao, no disclosures. Riyad Kherallah, no disclosures. Mackenzie Koetting, no disclosures. Leo Simpson, no disclosures. John Seger, consultant and speaker fees from Janssen Pharmaceuticals, Medtronic, Boehringer Ingelheim, Pfizer. Srikanth Koneru, no disclosures. Joseph Coselli, consulting and speaker fees from W. L. Gore & Associates, Bolton Medical Inc. Ourania Preventza, consulting fees from W. L. Gore & Associates. Vicente Orozco-Sevilla, none. Nastasya Manon, none. Guilherme V Silva, consulting and speaker fees from Medtronic, Abiomed, Ancora Heart.

## Abstract

**Background:** The safety and efficacy of leadless pacemakers (LP) in transcatheter aortic valve implant (TAVI) patients is not well known due to paucity of data. Herein, we compared outcomes between leadless pacemakers to traditional dual chamber pacemakers (DCP) following TAVI.

**Methods:** A single-center retrospective study was conducted, including a total of 27 patients with LP and 33 patients with DCP after TAVI between November 2013 to May 2021. We compared baseline demographics, pacemaker indications, percent pacing, ejection fractions, and pacemaker related complication rates.

**Results:** Leading indications for pacemaker implant were complete heart block (74% LP, 73% DCP) and high degree atrioventricular block (26% LP, 21% DCP). No significant differences were observed between LP and DCP in device usage and ejection fraction at 1, 6, and 12 months. Within each pacemaker group, we did not observe a significant reduction in percent ventricular pacing or ejection fraction at follow up. Three DCP patients required rehospitalization for pocket related complications.

**Conclusion:** From this single-center study, TAVI patients appear to have comparable pacemaker usage and ejection fraction between LP and DCP groups, suggesting that LP may be a reasonable alternative where single ventricular pacing is indicated. Larger studies are required to validate these findings.

## Introduction

Permanent pacemaker (PPM) implantation continues to be a leading complication following transcatheter aortic valve implant (TAVI).^1^ Nearly 1 in 10 patients receiving a TAVI will be expected to receive a PPM within 30 days of procedure.^2^ This population is expected to grow as TAVI volume continues to rise.^2-4^ Traditional PPMs are prone to developing pacemaker pocket hematomas, infections, and lead complications, with short term complication rates of up to 12%.^5^ In light of this growing pacemaker population and proven safety and reliability of leadless pacemakers (LP),^6,7^ there is a great interest in shifting from use of traditional PPM to leadless pacemakers in TAVI patients.

Since the Federal Drug Administration approval in 2016, the femoral inserted fully implantable leadless pacemaker has been a game changer by eliminating surgical pocket and lead related complications associated with traditional pacemakers.^8^ Such features confer multiple benefits to TAVI patients. The TAVI population is at high bleeding risk due to high prevalence (16-51%) of atrial fibrillation and anticoagulant usage.^9^ Thus, minimizing additional sources of bleeding leading to reinterventions would be of great benefit in this frail population. Second, a simultaneous TAVI and LP implant approach can be employed given the readily available femoral access during TAVI. In patients who develop intraprocedural high-degree atrioventricular block (HAVB) and complete heart block (CHB), current guidelines and expert consensus recommend a 24-hour observation period with a temporary transvenous pacer prior to PPM implant.^10-12^ However, such practice may result in increased length of stay and predispose patients to complications associated with temporary transvenous pacing.^13^ A simultaneous implant approach may be an appropriate and cost-effective solution in subset of patients with intra-procedure HAVB/CHB where single chamber ventricular pacing is justified. Retrospective studies have shown that most (65-87%) of TAVI associated HAVB/CHB occur intra-procedure^14,15^ and that majority (75%) of these patients ultimately require a PPM at discharge.^16^ Thus, a simultaneous implant approach may save this subset of patients from undergoing a separate procedure and allow for a shorter length of stay.^17^

Currently, clinical outcomes of leadless pacemakers in TAVI remains largely unknown due to scarcity of data, which are limited to case reports^18-21^ and one case series.^22^ Herein, we provide a single center experience comparing pacing and ejection fraction outcomes of TAVI patients with leadless and dual-chamber pacemakers.

## Materials and Methods

### Study population

A single center retrospective study was designed and approved by the local institutional review board. Patients who underwent leadless pacemaker implant following TAVI were identified from April 2018 to May 2021. All patients who received a LP met guideline indications^10,12,23^ for pacemaker implant and single ventricular chamber pacing, as determined by a consulting board-certified electrophysiologist. A total of 27 patients were identified, all of whom received a leadless Micra Transcatheter Pacemaker System (Medtronic, Minneapolis, MN, USA) in the septal apex location. For the comparison group, total of 33 patients who underwent dual chamber pacemaker implant following TAVI were identified between November 2013 to December 2020 from the local TAVI database of 443 patients.

### Data collection

Baseline demographics, medical comorbidities, conduction abnormalities, ejection fraction, valve type, and use of pre- and post-deployment balloon aortic valvuloplasty were collected via a standardized data extraction sheet. Procedural variables recorded include indication for intra-procedural (before patient left procedure room) pacemaker implant and indication for post-procedural (after patient left procedure room) pacemaker implant, time to pacemaker implant, fluoroscopy time, and length of stay. Pacemaker interrogation, including ventricular pacing percentage, capture threshold, pulse width, pacing impedance, R-wave amplitude, was obtained from clinicals at discharge, 1 month, 6 months, and 12 months follow up. Left ventricular ejection fraction (LVEF) was obtained from transthoracic echocardiography studies using the Simpson’s biplane method at baseline, 1 month, 6 months, and 12 months^24^. Post-pacemaker complications collected include pacemaker pocket hematoma requiring evaluation, pacemaker lead and generator extraction, pocket infection, and lead dislodgements. Procedure related complications were defined according to the Valve Academic Research Consortium-2 criteria.^25^ Six-month readmission rate and mortality at last follow up were recorded.

The diagnosis of cardiac conduction abnormalities warranting pacemaker implant was based on the definitions recommended by the American College of Cardiology, American Heart Association, and the Heart Rhythm Society guideline.^23^ High-degree atrioventricular block (HAVB) was defined as any of the following: second-degree AV block type 2 (Mobitz II) in the presence of a QRS at least 120 ms; 2:1 AV block in the presence of a QRS at least 120 ms; at least 2 consecutive P waves at a constant physiologic rate that do not conduct to the ventricles; transient third-degree AV block; in the setting of AF, a prolonged pause (>3 s) or a fixed slow (<50 beats/min) ventricular response rate. Complete heart block (CHB) or a third-degree atrioventricular block was defined as any of the following: P waves with a constant rate with dissociated ventricular rhythm (no association between P waves and R waves) or fixed slow ventricular rhythm in the presence of atrial fibrillation. The diagnosis of intraprocedural HAVB and CHB was made by continuous electrocardiogram monitoring in the procedure room. Persistent HAVB or CHB was defined as any episode that occurs during the procedure and either persists at the end of the procedure or lasts at least 15 minutes. Transient HAVB or CHB was defined as any episode occurring during the procedure that is transient and resolved at end of the procedure.

### Statistical analysis

We summarized continuous variables including pacemaker interrogation and LVEF as median (interquartile range [IQR]). We employed two-tailed nonparametric Mann-Whitney U test to assess differences in continuous variables between leadless pacemaker and dual-chamber pacemaker group. A one-tailed Mann-Whitney U test was used to assess whether pacemaker usage and LVEF decreased during follow up compared to baseline. Categorical variables were assessed for differences between groups via fisher’s exact test with alpha as 0.05.

## Results

A total of 27 subjects were included in the LP group and 33 subjects were included in the DCP group (table 1). Median age was 78 years for LP and 79 years for DCP group. A higher prevalence of baseline first-degree atrioventricular block in the DCP group (24% DCP vs 4% LP) was observed. There were no other significant differences in baseline conduction abnormalities. Common pre-existing abnormalities included atrial fibrillation (25.9% LP, 36.4% DCP) and right bundle branch block (22.2% LP, 27.3% DCP). There was no significant difference in baseline LVEF (57.5% LP, 60.0% DCP). Most patients received CoreValves (96.3% LP, 97% DCP) and there were no significant differences in use of pre and post-deployment aortic valvuloplasty.

**Table 1.** Baseline Characteristics.

|  | **Leadless**<br>**pacemaker**<br>**(n = 27)** | **Dual chamber**<br>**pacemaker**<br>**(n = 33)** | **p-value** |
| --- | --- | --- | --- |
| Age, y, median (IQR) | 78 (72-86) | 79 (70-84) | 0.77 |
| Female, no. (%) | 12 (44.4) | 23 (69.7) | ns |
| Body mass index, median (IQR) | 26.3 (23.9-32.2) | 29.2 (26.5-35.1) | 0.33 |
| Congestive heart failure, no. (%) | 10 (37.0) | 18 (54.5) | ns |
| Coronary artery disease, no. (%) | 17 (63.0) | 30 (90.1) | 0.01 |
| Hypertension, no. (%) | 22 (81.5) | 20 (60.6) | ns |
| Previous myocardial infarction, no. (%) | 3 (11.1) | 1 (3.0) | ns |
| Diabetes mellitus, no. (%) | 9 (33.3) | 16 (48.5) | ns |
| COPD, no. (%) | 5 (18.5) | 14 (42.4) | ns |
| Cerebral vascular disease, no. (%) | 3 (11.1) | 1 (3.0) | ns |
| GFR < 60 ml/min/1.73 m<sup>2</sup>, no. (%) | 8 (29.6) | 18 (54.5) | ns |
| Left ventricular ejection fraction, %, median (IQR) | 57.5 (47.0-66.1) | 60.0 (55.0-60.0) | 0.66 |
| Baseline conduction abnormalities |  |  |  |
| Atrial fibrillation, no. (%) | 7 (25.9) | 12 (36.4) | ns |
| First-degree atrioventricular block, no. (%) | 1 (3.7) | 8 (24.2) | 0.03 |
| Left anterior hemiblock, no. (%) | 3 (11.1) | 2 (6.1) | ns |
| Left bundle branch block, no. (%) | 0 (0.0) | 1 (3.0) | ns |
| Right bundle branch block, no. (%) | 6 (22.2) | 9 (27.3) | ns |
| Valve placed |  |  |  |
| Sapien, no. (%) | 0 (0.0) | 1 (3.0) | ns |
| Sapien 3, no. (%) | 1 (3.7) | 0 (0.0) | ns |
| CoreValve (1<sup>st</sup> gen), no. (%) | 0 (0.0) | 1 (3.0) | ns |
| CoreValve Evolut R (2<sup>nd</sup> gen), no. (%) | 4 (14.8) | 17 (51.5) | 0.006 |
| CoreValve Evolut Pro (3<sup>rd</sup> gen), no. (%) | 22 (81.5) | 14 (42.4) | 0.003 |
| Pre-deployment aortic valvuloplasty | 11 (40.7) | 9 (27.3) | ns |
| Post-deployment aortic valvuloplasty | 12 (44.4) | 12 (36.4) | ns |
| Abbreviations: COPD, chronic obstructive pulmonary disease; GFR, glomerular filtrate rate; IQR, interquartile range; ns, not statistically significant by fisher's exact test. |  |  |  |

There were no significant differences in pacemaker indication between two groups (table 2). Complete heart block was the leading indication for pacemaker implant during TAVI (63% LP, 47% DCP) and before discharge (74% LP, 73% DCP). Both populations had high rates of pacemaker implant during TAVI (70% LP, 62% DCP) and there was no significant difference in time to pacemaker implant between groups (median: 0 days for both groups). No difference in length of stay was observed (median: 8 days LP, 7 days DCP).

**Table 2.** Pacemaker Procedural Outcomes.

| **Outcomes** | **Leadless pacemaker**<br>**(n = 27)** | **Dual chamber**<br>**pacemaker**<br>**(n = 33)** | **p-value** |
| --- | --- | --- | --- |
| Intra-procedural pacemaker indication, no. (%) | 19 (70.3) | 21 (63.6) | ns |
| Persistent complete heart block | 15 (55.6) | 16 (48.5) | ns |
| Transient complete heart block | 2 (7.4) | 0 (0.0) | ns |
| Persistent high degree atrioventricular block | 1 (3.7) | 5 (15.2) | ns |
| Transient high degree atrioventricular block | 1 (3.7) | 0 (0.0) | ns |
| Post-procedural pacemaker indication, no. (%) | 8 (29.6) | 12 (36.4) | ns |
| Left bundle branch block | 0 (0.0) | 1 (3.0) | ns |
| High degree atrioventricular block | 5 (18.5) | 2 (6.1) | ns |
| Complete heart block | 3 (11.1) | 8 (24.2) | ns |
| Symptomatic bradycardia | 0 (0.0) | 1 (3.0) | ns |
| Time to implant, days, median (IQR) | 0 (0-1) | 0 (0-2) | 0.36 |
| Fluoroscopy time, mins, median (IQR) | 10.0 (7.0-12.9),<br>n = 15 | 10.3 (5.9-12.7),<br>n = 12 | 0.75 |
| Temporary transvenous pacemaker, no. (%) | 6 (22.2%) | 9 (27.3%) | ns |
| Length of stay, days, median (IQR) | 8 (4-15) | 7 (4-14) | 0.89 |
| Abbreviations: IQR, interquartile range; ns, not statistically significant by fisher's exact test. |  |  |  |

Median pacemaker follow up was 6 months in LP group and 1 month in DCP (table 3). There was no significant difference in pacemaker usage between groups at discharge (median: 72.9% LP, 99.0% DCP), 1 month (median: 26.1% LP, 90.2% DCP), 6 months (median: 42.3% LP, 84.0% DCP), and 12 months (median: 2.1% LP, 7.0% DCP). Within each cohort, ventricular pacing percentage varied widely and no significant reduction in ventricular pacing at 1, 6, and 12 months was observed. Median echocardiography follow up was 2 months in LP group and 6 months in DCP. There was no significant difference in LVEF at baseline, 1, 6, and 12 month follow up between groups. Compared to 1 month, there was also no statistically significant drop in ejection fraction at 6 and 12 month follow up within each cohort.

**Table 3.**
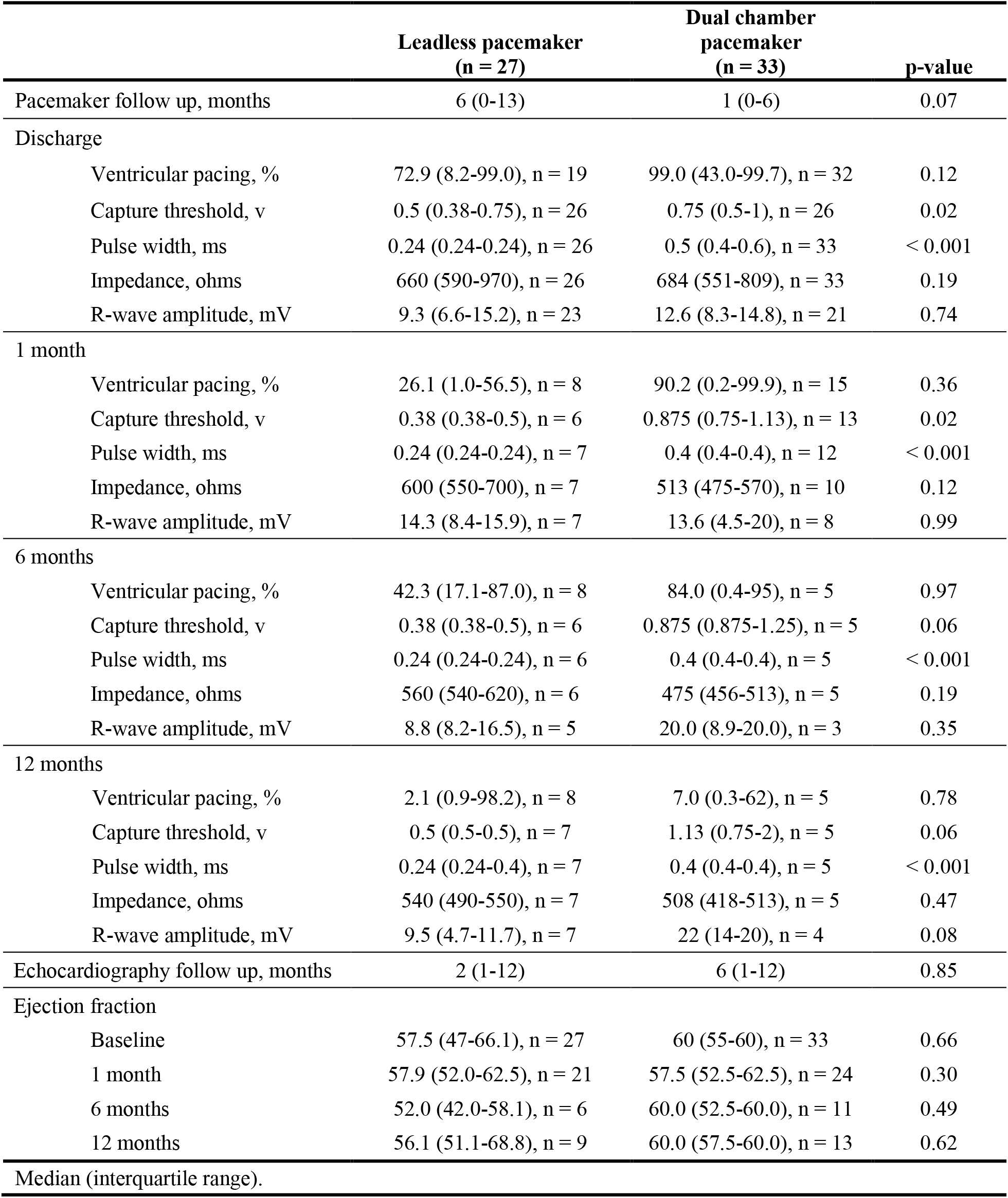
Pacemaker and Echocardiography Outcomes.

Three DCP patients required rehospitalization with in first 6 months due to pacemaker complications (table 4). Two patients required lead and generator extraction due to pacemaker pocket infection. One patient required pocket hematoma evacuation. Zero LP patients were re-hospitalized for pacemaker related complications. There was zero lead dislodgement or loss of capture events in the LP group while two patients in the DCP group experienced lead dislodgement requiring reintervention prior to discharge. Despite the larger femoral venotomy required to deliver the Micra leadless pacemaker system, zero minor vascular access complications were observed. Greater mortality was observed LP group (22.2%) compared to DCP group (9.1%).

**Table 4.** Post-Pacemaker Complications.

|  | **Leadless pacemaker**<br>**(n = 27)** | **Dual chamber**<br>**pacemaker**<br>**(n = 33)** |
| --- | --- | --- |
| Pocket hematoma evacuation, no. (%) | 0 (0.0) | 1 (3.0) |
| Lead and generator extraction, no. (%) | 0 (0.0) | 2 (6.1) |
| Pocket infection, no. (%) | 0 (0.0) | 3 (9.1) |
| Lead dislodgement, no. (%) | 0 (0.0) | 2 (6.1) |
| Minor bleeding, no. (%) | 1 (3.7) | 3 (9.1) |
| Minor vascular access complication, no. (%) | 0 (0.0) | 2 (6.1) |
| Acute kidney injury stage 1, no. (%) | 4 (14.8) | 2 (6.1) |
| Acute kidney injury stage 2, no. (%) | 1 (3.7) | 1 (3.0) |
| Rehospitalization, no. (%) |  |  |
| Pacemaker related | 0 (0.0) | 3 (9.1) |
| Not pacemaker related | 4 (14.8) | 10 (30.3) |
| Mortality, no. (%) |  |  |
| Pacemaker related | 0 (0.0) | 0 (0.0) |
| Not pacemaker related | 6 (22.2) | 3 (9.1) |
| Abbreviation: Minor bleeding and minor vascular access complication are defined by the Valve Academic Research Consortium-2 consensus. Acute Kidney Injury staging as defined by Acute Kidney Injury Network. |  |  |

## Discussion

In light of the continued high need of pacemaker implants following TAVIs and the validated safety of new leadless pacemakers in non-TAVI patients, a larger study of LP following TAVI is essential in understanding pacemaker performance and complications in this population. This study, to the authors’ knowledge, is the largest report of leadless pacemakers following TAVI along with the first study to compare outcomes to dual chamber pacemakers. Our study shows that in two TAVI cohorts of similar baseline characteristics, the performance of leadless pacemakers is comparable to that of dual chamber pacemakers with fewer complication rates.

Our study also validated the utility of a simultaneous LP implant approach for TAVI patients who develop intra-procedural conduction abnormalities. Our study observed a high rate DCPs placed during the TAVI procedure, which is a cumbersome process requiring switching from femoral to subclavian access mid procedure. Thus, employing a LP would greatly reduce the frequency of access site changes.

The cumulative percent time spent in ventricular pacing has been an area of intense interest in TAVI patients. The Dual Chamber and VVI Implantable Defibrillator (DAVID) trial, Multicenter Automatic Defibrillator Implantation Trial II (MADIT-II) trial, and Mode Selection Trial in Sinus Node Dysfunction (MOST) trial originally demonstrated that excessive pacing may pose deleterious effects on heart failure, hospitalizations, risk for atrial fibrillation and mortality.^26-28^ Specifically, right ventricular pacing over 40-50% of the time was associated with increased risk of worsening heart failure and hospitalizations.^27,28^ With our study, median pacing percentage was highly variable, ranging from 2% to 42% in the LP group and 7% to 90% in DCP group. Interpretation of our results is difficult in the context of the aforementioned studies since they were conducted nearly 20 years ago on non-TAVI patients. Additionally, recent studies have shown that pacemaker usage in TAVI patients is not only highly variable between individuals^29-31^ but also highly dynamic within each individual.^31^ Further supporting the safety of pacemaker usage in TAVI patients, evidence from systematic reviews have shown that pacemaker placement was not associated with increased mortality in mid to long-term TAVI studies.^32,33^ Thus despite our varied pacing percentages, there is low level of evidence to support that either patient cohorts was at significantly risk for adverse events from high pacing usage.

Pacemaker dependency is another metric of interest in TAVI patients. However, limitations exist based on how this outcome is defined and how data are collected. First, there is no consensus definition of “pacemaker dependency”. Small observational studies have reported pacemaker dependency rates of 40-60% during short to mid-term follow-up of TAVI patients with pacemaker implants for high degree or 3^rd^ degree heart blocks.^30,34,35^ However these studies are limited due to the reliance of a one-time spot follow-up electrocardiogram (ECG) as the sole method of assessing for pacemaker dependency. Such spot check technique can easily misclassify patients who require pacing during other times of the week as “not pacemaker dependent” and does not give a general representation of mean pacemaker usage. One study deserving special notice is by Meduri et al 2019, which employed a standardized protocol to assess true pacemaker dependence of each patient.^31^ The protocol designated such that each pacemaker’s backup rate be turned down to 30 beats per minute and only patients whose underlying rate is 30 or became symptomatic are declared truly pacemaker dependent. These study authors concluded that in a group of TAVI patients with predominant PPM indication of high degree and third degree heart blocks—similar to our study population—that pacing dependency was dynamic and was not statistically different between 1 month after implant (43%) and 1 year later (50%).^31^ This finding supports the notion that in patients who receive PPM for HAVB or CHB, that their dependence remains largely unchanged, similar to our observed results.

One strength of our study is we sought to compare the effect of prolonged right ventricular pacing on drop in ejection fraction in TAVI patients between LP and DCP patients. It is well known that TAVI patients with PPM implant experience small reductions in LVEF compared to those without PPM.^33^ Further, it is recognized that leadless pacemakers also may lead to LVEF reduction in non-TAVI patients.^36^ The postulated mechanism is that the electrical and mechanical dyssynchrony from RV apical pacing results in less effective ventricular contractions and changes in coronary blood flow.^37^ Thus, the effect of LP on TAVI ejection fraction has not been explored. In our study, we did not detect a significant difference in LVEF between LP and DCP groups at all time points. To our knowledge, this is the first study to demonstrate this in TAVI patients and supports LP safety in this population.

Main limitations of our study include small sample size, retrospective study design, and incomplete follow up. The reported effect size of pacemaker associated LVEF reduction is already small (effect size ∼ 0.26),^38^ thus our study is likely underpowered to detect any smaller differences that may exist between LP and DCP groups. While the LP group did observe greater mortality (22%) at follow up, this data is inconclusive in a retrospective study with incomplete follow up between groups.

## Conclusion

This study is the largest study to date on LP implant following TAVI and the first to compare outcomes to DCPs. We conclude that the femoral inserted fully implantable LPs is a suitable alternative in TAVI patients. Our results suggest that in the TAVI population, LPs have similar pacing needs and effect on ejection fraction as DCPs without increasing complication rates. Future larger studies are required to investigate long-term outcomes in this population.

## Data Availability

All data produced in the present study are available upon reasonable request to the authors.

## Acknowledgements

none

## Author contributions

concept/design (all authors), data analysis/interpretation (FG, RK, GVS), drafting article (FG, RK, OP, GVS), critical revision of article (FG, RK, OP GVS), approval of article (all authors), statistics (FG, RK), data collection (FG, MK).

